# Shouldering our Way into a More Meaningful Research Agenda for Atraumatic Shoulder Pain: *A Priority Setting Study*

**DOI:** 10.1101/2024.08.22.24312355

**Authors:** KD Lyng, TK Børsting, MB Clausen, AH Larsen, B Liaghat, KG Ingwersen, M Bateman, A Rangan, KT Bjørnholdt, DH Christiansen, SL Jensen, JNL Thomsen, K Thorborg, C Ziegler, JL Olesen, MS Rathleff

**Author notes:** **Corresponding Author**, Kristian Damgaard Lyng, Department of Health Science and Technology, Aalborg University, Selma Lagerløfs Vej 249, 9260 Gistrup, Denmark.

## Abstract

**IMPORTANCE:** Atraumatic shoulder pain poses a significant burden to society and the individual. There is a growing need for involving patients and other stakeholders in setting the research agenda.

**OBJECTIVE:** To use the voice of people living with atraumatic shoulder pain, healthcare practitioners, and their relatives to establish research questions.

**DESIGN, SETTING, AND PARTICIPANTS:** This priority-setting study followed a modified approach originally formulated by the James Lind Alliance (JLA). The process consisted of six phases (initiation, consultation, collation, prioritization, validation, and reporting) and included two e-surveys and two separate virtual workshops. Data collection started on June 2021 until June 2023. We included people with atraumatic shoulder pain, relatives, healthcare practitioners managing shoulder pain, and researchers conducting research within the field.

**EXPOSURES:** The first e-survey included basic demographic questions and the possibility to submit at least one and a maximum of five potential research questions. Based on a thematic analysis, questions were arranged into themes and related questions. In the second e-survey, participants were asked to prioritize the questions. Finally, two priority-setting partnership workshop was used to formulate a top-10 list.

**MAIN OUTCOMES AND MEASURES:** A top-25 and top-10 list of research questions related to atraumatic shoulder pain.

**RESULTS:** Initially, 297 participants submitted 1080 potential research questions. In the second e-survey 290 participants prioritized these questions resulting in a compilation of the top 25. Based on discussions from the workshops with a total of 21 participants, a top 10 list was created.

**CONCLUSIONS AND RELEVANCE:** In the final top 10 list, the three research questions with the highest ranking concerned 1) translating the best available knowledge into clinical practice, 2) preventing shoulder pain, and 3) identifying who benefits from surgery. These questions informs future research funding and projects relating to atraumatic shoulder pain.

**Key points:** *Question:* What are the top research priorities with atraumatic shoulder pain that can help directing future research agendas for improving current care.

*Findings:* This priority-setting study included two e-surveys and two virtual workshops to establish top-10 research questions based on input from people living with atraumatic shoulder pain, healthcare practitioners, and relatives. The final list focused on improving knowledge translation to clinical practice, prevention of pain and identifying who benefits from surgery.

*Meaning:* This lists provide funders, policymakers, and researchers with user-generated knowledge on where to prioritize their resources for creating meaningful improvements in the management of atraumatic shoulder pain.

## Introduction

The prevalence of shoulder pain increases with age and is one of the most common reasons people seek care for pain in the upper extremities.^1^ Atraumatic shoulder pain is a term which covers multiple specific diagnoses, such as subacromial pain syndrome, rotator cuff tendinopathy, adhesive capsulitis, arthritis, degenerative rotator cuff tears, and instability.^2^ There is not yet a universal agreement of terminology or clear aetiology, which hampers cross-study comparison.^3^ Also, there is a lack of association between structural changes and pain, highlighting the need to focus on patients’ experiences and symptoms.^4–6^

Atraumatic shoulder pain poses a substantial burden both for the individual and the society.^7–11^ Recent research shows that 70% of the economic impact associated with shoulder disorders in the working-age population is attributed to sick leave.^11^ National cost-of-illness studies further highlight that the economic burden is attributed to 20% of the patients, who account for 66% of the total cost of the healthcare system.^11^ The economic burden of shoulder pain has brought an emphasis on the healthcare costs associated with the condition and its various treatments^12–15^. Costs may be a high priority for some policymakers; however, costs are unlikely to be the main concern for people with lived experience of shoulder pain. Despite large research efforts and advancement, this issue is further highlighted by the existing uncertainties in the treatment of atraumatic shoulder pain. Recent insights from large trials like the SExSI, GRASP, UK-FROST, and CSAW trials have established that no specific treatment strategy has shown superiority for common atraumatic shoulder conditions.^13–16^

Despite the recent development of evidence-based interventions, a significant proportion continues to experience pain.^15–17^ This underlines the need for further research to optimize the management of atraumatic shoulder pain and to identify what is important to end-users dealing with atraumatic shoulder pain using the safest and most cost-effective approach.

Within recent years, there has been a growing interest in patient involvement in healthcare research, especially in musculoskeletal pain research across all ages.^18–21^ Several national and international initiatives have been launched to both involve patients and other stakeholders in research and also to provide guidance to researchers working towards patient and public involvement in research.^22–25^ As recently highlighted in two *Lancet Rheumatology* articles, there is a great need to identify and combat societal issues through the active involvement of patients and the public in research.^26,27^

Several different methodologies and frameworks have been used for the inclusion of various stakeholders in general musculoskeletal pain research prioritizing, including The Child Health and Nutrition Research Initiative (CHNRI) and James Lind Alliance Priority Setting Partnerships (JLA-PSPs).^28,29^ While one surgery-specific shoulder priority-setting study and priority-setting studies on the broader musculoskeletal pain field have already been conducted, it is unknown whether research priorities differ when addressing the entire health situation and management of people living with atraumatic shoulder pain.^20,21,30–33^

Adhering to the JLA-PSP principles, this study aimed to establish research priorities for the field of atraumatic shoulder pain by involving people with atraumatic shoulder pain, their carers, healthcare professionals, and researchers.

## Methods

### Study Design

The reporting of this study is guided by the Guidance for Reporting Involvement of Patients and the Public (GRIPP2) short-form checklist and the Reporting Guideline for Priority Setting of Health Research (REPRISE).^34^ The study was conducted using methods similar to those previously described.^18,21^ The study used a modified approach adapted from the James Lind Alliance guidance for conducting Priority Setting Partnerships.^29^ This approach has previously been used in other pain conditions to identify future patient-oriented research priorities.^18,21,31,35,36^ The process consisted of six phases (initiation, consultation, collation, prioritization, validation, and reporting) (See **Figure 1** for an overview of all phases). All data were processed and stored in accordance with the General Data Protection Regulation (GDPR). Regional ethical approval was deemed exempt due to the low-risk nature of the study. The study was conducted between June 2021 and February 2023. All data were collected in Denmark through e-surveys in the secure web-based software REDcap (Research Electronic Data Capture) hosted at Aalborg University.^37,38^

**FIGURE 1.** Overview of Study Phases 1-6.

### Search strategy and selection criteria

To ensure relevancy of the project and to inform the findings of the study, we searched PubMed and Embase from Jan 1, 2003 to December 1, 2023 for peer-reviewed English-language articles. The search strategy was separated into two strategies. The first included search terms related to atraumatic shoulder pain. For this search strategy the terms included “shoulder*”, “shoulder pain”, “atraumatic shoulder pain”, “non-traumatic shoulder pain” or “non-specific shoulder pain” combined with “epidemiology”, “diagnosis”, “management”, “therapy” and “treatment”. The second search was concerning chronic musculoskeletal pain and participatory research. This search strategy included search terms such as “chronic musculoskeletal pain”, “adults”, “management”, “therapy” and “treatment” combined with “participatory research”. All articles were selected based on their data and relevance for this study. Studies using primary data or high-quality systematic reviews were prioritised. Furthermore, if two or more studies existed concerning similar aims, the most recent study was prioritised.

### Context and Scope

This priority-setting study was conducted across all regions of Denmark, anchored in Aalborg at Aalborg University. The project was initiated as a part of a continuous effort to identify research priorities across musculoskeletal pain conditions.^21,39^ The outcome of this study primarily targets researchers, policymakers, funders, and the industry, who all have an interest in improving the lives of people living with atraumatic shoulder pain.

### Initiation

In the initiation phase, the steering group (all authors) discussed the scope of the project and designed the protocol. Further, different stakeholders were invited to the initiation of the study, including patient organizations, patients, and carers, all of whom provided feedback on the scope of the study and the protocol. Stakeholders with various backgrounds (e.g., gender, age, work situation, ethnicity, and geographical location) were purposively sampled to ensure representativeness and provide a voice for all relevant stakeholders. All stakeholders were recruited through e-mail.

### Consultation

This step involved gathering research questions directly from all relevant stakeholders in a Danish context. To accommodate this, the steering group developed an electronic survey (e-survey), which was tailored to fit each respondent category (i.e., people living with atraumatic shoulder pain, carers, and healthcare practitioners). The survey consisted of various demographical questions and the opportunity to state research questions (“What do you think future research should prioritize?”) for future research in relation to the management of atraumatic shoulder pain. The e-survey was piloted on five people with atraumatic shoulder pain, five healthcare practitioners working with people with atraumatic shoulder pain, and three carers. The participants from the pilot tests were asked to provide feedback on the wording and the appropriateness of the e-survey. The survey used a multimodal recruitment process. This involved distribution through newsletters from patient organizations, personal and professional networks, and targeted advertisement through Facebook and LinkedIn. No reimbursement was given for participation. All participants were aged 18 years or above and were residents of Denmark. Three groups were recruited for the study:

**1)** People with atraumatic shoulder pain, their carers, or relatives
**2)** Healthcare practitioners with clinical experience in managing shoulder pain
**3)** Researchers with experience in conducting research on shoulder pain

People with atraumatic shoulder pain were included if they had experienced consistent non-traumatic shoulder pain for more than three months. Only authorized healthcare practitioners, such as medical doctors, psychologists, physiotherapists, and chiropractors, were included in group 2. All participants who completed the survey were invited to take part in the later phases of the study.

### Collation

After the collection of research questions from Step 1, a thematic analysis was conducted in agreement with the James Lind Alliance guidebook, through a thematic text analysis as described by Braun and Clarke.^29,40^ This involved the following steps: familiarisation with data, coding of data, searching for themes, reviewing themes, defining, and naming themes, and presenting the results. KDL and TKB familiarised themselves with the raw data through naïve reading. After this, both researchers independently coded each individual research question and categorized potential domains. All potential domains were organized in a mind map for vertical and horizontal interpretation. This was then condensed into a table including the main themes, sub-themes, sub-sub-themes, indicative questions, examples, and summative descriptions. The thematic analysis was presented to two people with atraumatic shoulder pain and two healthcare practitioners, who provided feedback on the thematic analysis to ensure that it was kept true to the data. Furthermore, they validated the excluded inquiries (e.g., answers that were not relevant to atraumatic shoulder pain and unreadable inquiries) and determined whether these research questions were conclusively irrelevant or not. All individuals were recruited through patient organizations and professional networks. NVivo 12 (NVivo qualitative data analysis software; QSR International Pty Ltd. Version 12, 2018) was used for the thematic analysis.

### Prioritization

#### Interim prioritization

To reduce the list of research questions emerging from the previous steps, we developed an interim prioritization e-survey. The results from the thematic analysis were summarized into questions based on feedback from two people with atraumatic shoulder pain, one carer, and one healthcare practitioner. The e-survey was distributed through clinical practices across Denmark and social media (Facebook and LinkedIn) and to participants from the consultation phase who had consented to participate in later stages. In the interim prioritization e-survey, the respondents were asked to choose the ten most important research questions and the five least relevant research questions. The participants ranked the importance of each research question on a 5-point Likert scale from 1 to 5, with 1 = not at all important, two = low importance, 3 = neutral, 4 = important, and 5 = very important. An overall score for each question was determined by counting the number of times the research question was rated “very important” or “important”. The top 25 research questions were presented to the steering group and constituted the list of final questions for the workshops. The order of the top 25 research questions was randomized prior to the workshops to minimize the influence on the voting. Before completion of the prioritization exercise, the participants were again invited to take part in later phases of the study. The top 25 research questions from the interim prioritization exercise will be presented alongside the top 10 questions to provide full transparency on the weighting from each step and the methodological approach.

### Validation

#### Workshop to Determine the Top 10 Research Priorities

To determine the final top 10 of the research priorities established from the previous steps, we invited people with atraumatic shoulder pain, carers, and healthcare practitioners to participate in two separate workshops. To ensure that all voices were heard, we created two groups consisting of healthcare practitioners in one workshop (W1) and people with atraumatic shoulder pain and carers in another workshop (W2). The participants were recruited through previous e-surveys from the consultation and prioritization phase, clinical practices across Denmark, and social media (Facebook and LinkedIn). Steering group members with no conflict of interest were also invited to participate. The participants were purposively sampled to include participants with various backgrounds (e.g., ethnicity, education, work situation), diagnosis, care journey (i.e., healthcare practitioners seen), sex, age, and duration of symptoms (in months). We aimed to recruit an equal number of people with atraumatic shoulder pain, carers, and healthcare practitioners of various demographics to ensure representativeness. The first workshop (W1) was facilitated by three authors of the project (KDL, TKB and AHL). The second workshop (W2) was facilitated by one author (KDL). Both workshops included both small and large group exercises. All participants received the list of the top 25 research questions and an introduction to the aim of the project prior to the workshop. The participants were asked to familiarize themselves with the top 25 research questions. The workshop was initiated with an introduction to the agenda, and all participants were informed about the previous steps of the process that led to the workshop. In each workshop, the participants were first divided into small groups and asked to identify the *least* important research questions. Then, the questions were removed based on consensus from all the attending participants. Again, the participants were divided into small groups and asked to select the *most* important research questions, which were discussed to obtain consensus in the entire group of participants. Using the nominal group technique, each participant was asked to forward the three most important research questions to the lead author (KDL). Using this approach minimized the influence that participants could potentially pose on each other. The answers from all participants were used to formulate the final top 10 list of research questions within the management of atraumatic shoulder pain. All votes were given a nominal value, and the total values were used to create the final top 10. All votes were weighed equally.

## Results

Of all the 212 included healthcare practitioners, 4 (2%) had three months – 1 year of experience working with the population, 10 (5%) had 1-2 years of experience, 19 (9%) had 2-5 years of experience, 30 (14%) had 5-10 years of experience, 85 (40%) had 10-20 years of experience, and 64 (30%) had +20 years of experience (**Table 1**). Of the total 385 included people with atraumatic shoulder pain, 69 (18%) had experienced atraumatic shoulder pain between 3 months – 1 year, 58 (15%) had 1-2 years of pain duration, 111 (29%) had 2-5 years of pain duration, 62 (16%) had 5-10 years of pain duration, 50 (13%) had 10-20 years of pain duration, and 35 (9%) had +20 years of pain duration. The most consulted practitioners were physiotherapists (339 (88.1%)) and general practitioners (338 (87.8%)). An orthopaedic surgeon had been seen by 209 (54.3%) of the patients, and a rheumatologist, chiropractor, psychologist, or neurologist had been seen by 169 (43.9%), 164 (42.6%), 57 (14.8%), and 44 (11.4%) respectively. Further, 26.5% of the patients had consulted a specialized pain clinic. In the first phase, 297 participants (177 (59.5%) females and 120 (40.5%) males) completed the e-survey and submitted 1080 potential research questions. Of these, 230 (77.5%) participants were people with atraumatic shoulder pain, one was a career (0.3%), and 66 (22%) were healthcare practitioners. The potential questions were condensed into 16 main themes (See **Panel 1**) and 94 subthemes that were formulated into research questions. In the interim prioritization exercise, 290 completed the e-survey, and 94 questions were reduced to 25 research questions for further use in the workshops (**Table 2**). The top three themes were treatment (prioritized by 211 (73%)), self-management (prioritized by 171 (59%)), and prognosis (prioritized by 153 (53%)). Eleven healthcare practitioners participated in the first virtual workshop (W1) to determine the top research priorities in relation to atraumatic shoulder pain. Three invited participants (one medical doctor, one psychologist, and one physiotherapist) did not attend the workshop because of unexpected work issues. The results from the nominal voting can be seen in **Table 3**. Eight people with atraumatic shoulder pain and two carers participated in the second workshop (W2), which also took place online. Four had been diagnosed with subacromial pain syndrome, two with glenohumeral osteoarthritis, one with adhesive capsulitis, and one with an atraumatic rotator cuff tear (supraspinatus). Nine invited participants (seven people with atraumatic shoulder pain and two carers) did not attend the workshop because of pain-related issues (n = 5) or unexpected work issues (n = 4).

**TABLE 1.** Participants’ Characteristics.

**TABLE 2.** Top 25 Research Questions based on Interim Prioritisation.

**TABLE 3.** Top 10 Research Question Priorities.

**PANEL 1.** Listing of Main Themes.

## Discussion

There is an increased need to involve end-users in research to maximize the impact of research for the benefit of society.^41^ We established the priorities for research in atraumatic shoulder pain. These findings can guide the future direction of research to address the end-user’s priorities and needs. The three research questions with the highest ranking in the top 25 list (based on 290 e-survey answers) included: 1) which exercise regimen is the most effective, 2) how can patients learn to self-manage their own pain, and 3) how effective is painful exercising compared with non-painful exercising. From the top 10 list (based on two workshops with 19 participants), the three research questions with the highest ranking were: 1) how can we improve the translation of research into clinical practice, 2) how can we prevent atraumatic shoulder pain, and 3) who benefits from surgery, and who does not? The results from this study represent different areas of interest across and outside the healthcare sector.

The research question with the highest ranking in the top 10 list is “*How can we improve the translation of research into clinical practice?”*. This priority shows that healthcare practitioners, people in pain, and careers have a priority to strengthen the implementation of the most up-to-date knowledge. No published studies have investigated how to translate research on atraumatic shoulder pain most effectively into clinical practice. Furthermore, only limited evidence exists regarding the most effective implementation strategy for new knowledge in terms of musculoskeletal pain.^42^ This underlines a difference between end-users’ priorities and the published literature. The priority of knowledge translation aligns with the policies of the World Health Organization (WHO) and the Evidence-Informed Policy Network (EVIPNet). These policies emphasize the need to accelerate the implementation of research into clinical practice and the healthcare system in general.^43,44^

Of the total 35 research priorities, 15 of these priorities from both the top 25 and 10 lists were related to improving the existing treatments and how to tailor the treatments to the individual. This aligns well with larger trials published in leading medical journals within the last few years.^13–16,45,46^ From the top 10 list, three priorities included traditional treatments, including exercise and surgery, which have been investigated intensively within the last decade.^15–17,47–52^ The majority of the research conducted on surgery for the most common shoulder pain conditions has failed to demonstrate a clear improvement compared to a placebo,. This has led to most guidelines advising strongly against surgery.^53,54^ Recent studies have shown a decrease in surgery rates during the last years. However, surgical procedures such as subacromial decompression are still frequently performed indicating a need for more knowledge on who might benefit from surgery and implementation of research into practice.^55,56^

Exercise only provides small to moderate effect sizes comparable to laser therapy and extracorporeal shock wave therapy after 3-6 months.^17^ Recent trials have underpinned the lack of superiority of exercise compared to best-practice advice^15^ (irrespective of the dosage of exercise).^16^ The priorities that concern exercise and surgery highlight the need for more research on how we can optimize health through a combination of treatments to maximize effectiveness. To the author’s knowledge, most of the existing randomized control trials have focused on single interventions or multimodal interventions with only limited capacity for individualization or person-centred care. Policymakers such as the International Consortium for Personalised Medicine (ICPerMed) highlights the increase in interest in order to build capacity for more personalized care in health care research.

We identified the need for more research on novel interventions, including shared decision-making, patient education, and self-management interventions for atraumatic shoulder pain. These findings underline the need for management strategies that focus on challenging common misconceptions and supporting patients’ agency, which has been underprioritised in current literature.^17^ Encouraging active patient involvement and agency has been recognized by global organizations such as the European Union and WHO. This further stress the importance of conducting more research empowering people living with atraumatic shoulder pain to gain more agency and learn more about their own condition.

Other key priorities for future research include a better understanding of disease development, prevention, and prediction of how atraumatic shoulder pain influence’s function, workability, and quality of life. These findings correspond to a previously published evidence and gap map concerning chronic musculoskeletal pain, in which a minority of the included systematic reviews (n=457) focused on outcomes such as work-related health and quality of life. Further, the findings highlighted the low quality of the existing evidence.^57^ The authors advocate for future evidence and gap-mapping studies that could systematically identify disparities between the current evidence and the research priorities derived from stakeholders for atraumatic shoulder pain. Potentially, the combined knowledge of such studies will help policymakers, decision-makers, and researchers determine where to invest their resources.

An important strength of our study is that we closely collaborated with end-users throughout the entire study process. This helped ensure that the final product was kept as relevant to the end-users as possible. Furthermore, the end-users helped decide what was relevant and what was irrelevant throughout, again strengthening the relevancy of this study and minimizing potential biases posed by the steering group. We were guided by the JLA-PSP approach, which follows a specific set of steps to capture research priorities and formulate top 10 lists. Capturing research uncertainties through several steps, and using several different methods strengthens the validity of our priority lists and creates a foundation for an equal and transparent discussion of future research priorities. Furthermore, we also present the results from both the e-surveys and the workshops to be fully transparent and to highlight the differences between the two steps. Based on previous experience and feedback, we decided to conduct two workshops to provide patients with a better environment to express their views on equal terms with other persons in the same situation. We recommend this for future studies capturing research priorities.

Our study is not without limitations. Establishing priorities using these methods lacks a deep understanding of the “why” of the proposed research questions. Our research questions should be interpreted as valuable insights into which areas matter to the stakeholders but not why they matter. We are not able to provide credible explanations for the underlying reasons and mechanisms for why these research priorities emerged from our end-users. The priorities could, in theory, have multiple meanings. As an example, the priority “who benefits from surgery” could also represent patients feeling unjustly denied surgery, patients operated but not experiencing benefit, or clinicians doubting correct indications for surgery. This illustrates the need for patient education, adjusting patient expectations, or large RCTs with subgroup analysis to establish core indications of a specific surgery. Future studies should explore the reasons behind specific research priorities and explore if some priorities emerge as a result of poor knowledge translation.

Several studies have identified research priorities for the general musculoskeletal pain field ^20,21,31,33^. In this study, we aimed to be more specific and focused on atraumatic shoulder pain to deliver more specific priorities. This may be important for future priority-setting studies to consider as research priorities vary across different healthcare-related contexts.^39^ Despite our narrower focus, atraumatic shoulder pain is still a broad term encompassing several heterogeneous diagnoses, which are often treated through different care pathways. Hence, some priorities may be more or less valid for individual diagnoses and should be interpreted with care. Creating actionable research priorities is a difficult task that requires the priority to be formulated as a testable research question while the intended meaning of the original priority remains.^58^ It is questionable if all the generated research priorities can be translated into a falsifiable hypothesis that can be tested through traditional research methodologies. Importantly, research priorities also provide useful insight to policymakers and funders of research as the priorities provide information on what stakeholders jointly report as relevant for improving certain research areas. Therefore, we recommend that future research projects informed by the outcomes of this study formulate their research questions in close collaboration with relevant patient partners and healthcare practitioners.

Our study had an overrepresentation of people in pain (versus healthcare practitioners/careers) throughout all phases, predominantly with a Danish background. While this limits the applicability and generalisability to other groups, countries, healthcare systems and questions whether we have captured all research priorities, it also underpins that there is a need to create effective recruitment strategies for ethnic minorities and other underrepresented groups across society. Lastly, while several systematic steps are taken to obtain the top priority list, these priority-setting studies are not inherently protected against potential biases or agendas of the participants, including secondary gain issues. This is important when interpreting our findings. Future studies should also consider refining and validating research priorities from people with different lived experiences rather than specific diagnoses and in different life situations such as litigation or awaiting disability payments.

In conclusion, our study adds crucial knowledge to shape the future research agenda within atraumatic shoulder pain. Our study finds that priorities related to which exercise regimen is the most effective and how we can most effectively translate research into clinical practice are the two research questions with the highest ranking across the top 25 and top 10 lists, respectively. Funders and researchers should consider focusing on the priorities derived from this study to inform proper allocation of funds and resources to meet the priorities of people living with atraumatic shoulder pain and other end-users. Creating more end-user-driven innovation and research is highly needed within this area, and this study will provide guidance for accommodating this need identified by people living with atraumatic shoulder pain.

## Author Contributions

KDL, MBC, and MSR drafted the first version of the manuscript. KDL and TKB created all surveys with feedback from the rest of the author group. Data from the consultation and prioritization phase were collected, analyzed, and summarised by KDL and TKB. MBC, MSR, and CZ contributed to the analysis. CZ was in charge of reviewing the entire data analysis and ensuring the perspectives of the stakeholders. KDL, TKB, and AHL were in charge of the workshops, including setup, data collection, analysis, and summarizing. MB and AR provided significant expert methodological considerations throughout the entire process. KDL, TKB, MBC, AHL, BL, MB, AR, KTB, DHC, SLJ, JNLT, KT, CZ, JLO, and MSR all helped support the recruitment for all phases. KDL, TKB, MBC, AHL, BL, MB, AR, KTB, DHC, SLJ, JNLT, KT, CZ, JLO, and MSR made significant contributions to the design, interpretation, approval of data analysis, reporting (i.e., manuscript writing) and revisions.

## Supporting information

Tables and panel

## Data Availability

All data produced in the present study are available upon reasonable request to the authors.

## Acknowledgements

The authors would like to thank all the participants throughout all phases. We are grateful for your input, and we are honoured to act as the conveyor of all your voices. Furthermore, we want to thank the stakeholder group for providing essential input during the initiation of the study and the interim prioritization.

## Informed Consent

Informed consent was collected to ensure that participants accepted that we could contact them at later stages of the project and that data from this study were stored on a secure file share.

## References

1 Lucas J, Doorn P van, Hegedus E, Lewis J, Windt D van der. A systematic review of the global prevalence and incidence of shoulder pain. BMC Musculoskelet Disord 2022; 23: 1073.

2 Lee DYL, Haas R, Wallis JA, O’Connor DA, Buchbinder R. Clinical practice guidelines for the management of atraumatic shoulder conditions: protocol for a systematic review. Bmj Open 2021; 11: e048297.

3 Witten A, Mikkelsen K, Mayntzhusen TW, et al. Terminology and diagnostic criteria used in studies investigating patients with subacromial pain syndrome from 1972 to 2019: a scoping review. Br J Sports Med 2023; 57: 864–71.

4 Harris JD, Pedroza A, Jones GL, et al. Predictors of Pain and Function in Patients With Symptomatic, Atraumatic Full-Thickness Rotator Cuff Tears. Am J Sports Med 2012; 40: 359–66.

5 Dunn WR, Kuhn JE, Sanders R, et al. Symptoms of Pain Do Not Correlate with Rotator Cuff Tear Severity. *J Bone Jt Surg*, Inc 2014; 96: 793–800.

6 Curry EJ, Matzkin EE, Dong Y, Higgins LD, Katz JN, Jain NB. Structural Characteristics Are Not Associated With Pain and Function in Rotator Cuff Tears. Orthop J Sports Med 2015; 3: 2325967115584596.

7 Kuijpers T, Tulder MW van, Heijden GJ van der, Bouter LM, Windt DA van der. Costs of shoulder pain in primary care consulters: a prospective cohort study in The Netherlands. Bmc Musculoskelet Di 2006; 7: 83.

8 Aurora A, McCarron J, Iannotti JP, Derwin K. Commercially available extracellular matrix materials for rotator cuff repairs: State of the art and future trends. J Shoulder Elb Surg 2007; 16: S171–8.

9 Virta L, Joranger P, Brox JI, Eriksson R. Costs of shoulder pain and resource use in primary health care: a cost-of-illness study in Sweden. Bmc Musculoskelet Di 2012; 13: 17– 17.

10 Marks D, Comans T, Bisset L, Thomas M, Scuffham PA. Shoulder pain cost-of-illness in patients referred for public orthopaedic care in Australia. Aust Health Rev 2019; 43: 540–8.

11 Sørensen L, Tulder M van, Johannsen HV, et al. Costs of shoulder disorders in Denmark; a nationwide cost-off-illness study investigating 617,334 patients and matched controls. Pain 2022; Publish Ahead of Print. DOI:10.1097/j.pain.0000000000002610.

12 Carr A, Cooper C, Campbell MK, et al. Effectiveness of open and arthroscopic rotator cuff repair (UKUFF): a randomised controlled trial. Bone Jt J 2017; 99-B: 107–15.

13 Beard DJ, Rees JL, Cook JA, et al. Arthroscopic subacromial decompression for subacromial shoulder pain (CSAW): a multicentre, pragmatic, parallel group, placebo-controlled, three-group, randomised surgical trial. Lancet Lond Engl 2018; 391: 329–38.

14 Rangan A, Brealey SD, Keding A, et al. Management of adults with primary frozen shoulder in secondary care (UK FROST): a multicentre, pragmatic, three-arm, superiority randomised clinical trial. Lancet 2020; 396: 977–89.

15 Hopewell S, Keene DJ, Marian IR, et al. Progressive exercise compared with best practice advice, with or without corticosteroid injection, for the treatment of patients with rotator cuff disorders (GRASP): a multicentre, pragmatic, 2 × 2 factorial, randomised controlled trial. Lancet 2021; 398: 416–28.

16 Clausen MB, Hölmich P, Rathleff M, et al. Effectiveness of Adding a Large Dose of Shoulder Strengthening to Current Nonoperative Care for Subacromial Impingement: A Pragmatic, Double-Blind Randomized Controlled Trial (SExSI Trial). Am J Sports Medicine 2021; 49: 3040–9.

17 Babatunde OO, Ensor J, Littlewood C, et al. Comparative effectiveness of treatment options for subacromial shoulder conditions: a systematic review and network meta-analysis. Ther Adv Musculoskelet Dis 2021; 13: 1759720X2110375.

18 Birnie KA, Dib K, Ouellette C, et al. Partnering For Pain: a Priority Setting Partnership to identify patient-oriented research priorities for pediatric chronic pain in Canada. Cmaj Open 2019; 7: E654–64.

19 Beresford P, Russo J. Patient and public involvement in research. In: Anell A, Nolte E, Merkur S, editors. Achieving Person-Centred Health Systems: Evidence, Strategies and Challenges. European Observatory on Health Systems and Policies. Cambridge: Cambridge University Press; 2020. p. 145–72. 2020; : 145–72.

20 Paskins Z, Farmer CE, Manning F, et al. Research priorities to reduce the impact of musculoskeletal disorders: a priority setting exercise with the child health and nutrition research initiative method. Lancet Rheumatology 2022; 4: e635–45.

21 Lyng KD, Larsen JB, Birnie KA, et al. Participatory research: a Priority Setting Partnership for chronic musculoskeletal pain in Denmark. Scand J Pain 2022; 0. DOI:10.1515/sjpain-2022-0019.

22 European Patient’s Forum. European Patient’s Forum—The Value+Handbook 2022. https://www.eu-patient.eu/.

23 Patient Centered Outcomes Research Institute. 2022. https://www.pcori.org.

24 National Institute for Health and Care Research. 2022. https://www.nihr.ac.uk/.

25 International Collaboration for participatory Health research. International Collaboration for participatory Health research, the Canadian Institutes for Health Research 2022. http://www.icphr.org/.

26 Yeoh S-A, Burke B, Castelino M, et al. Patient and public involvement in rheumatology research: embracing the wave of change. Lancet Rheumatology 2021; 3: e540–2.

27 Berkovic D, Ackerman I, Buchbinder R. Patient-led research in rheumatology: the way forward? Lancet Rheumatology 2023; 5: e180.

28 Rudan I, Gibson JL, Ameratunga S, et al. Setting Priorities in Global Child Health Research Investments: Guidelines for Implementation of the CHNRI Method. Croat Méd J 2008; 49: 720–33.

29 Alliance JL. National Institute for health research, the James Lind alliance Guidebook: version 10. 2021.

30 Foster NE, Dziedzic KS, Windt DA van der, Fritz JM, Hay EM. Research priorities for non-pharmacological therapies for common musculoskeletal problems: nationally and internationally agreed recommendations. Bmc Musculoskelet Di 2009; 10: 3.

31 Rangan A, Upadhaya S, Regan S, Toye F, Rees JL. Research priorities for shoulder surgery: results of the 2015 James Lind Alliance patient and clinician priority setting partnership. Bmj Open 2016; 6: e010412.

32 Beneciuk JM, Verstandig D, Taylor C, et al. Musculoskeletal pain stakeholder engagement and partnership development: determining patient-centered research priorities. *Res Involv Engagem* 2020; 6: 28.

33 Slater H, Jordan JE, O’Sullivan PB, et al. “Listen to me, learn from me”: a priority setting partnership for shaping interdisciplinary pain training to strengthen chronic pain care. Pain 2022; Publish Ahead of Print. DOI:10.1097/j.pain.0000000000002647.

34 Tong A, Synnot A, Crowe S, et al. Reporting guideline for priority setting of health research (REPRISE). BMC Méd Res Methodol 2019; 19: 243.

35 Poulin P, Shergill Y, Romanow H, et al. Researching what matters to improve chronic pain care in Canada: A priority-setting partnership process to support patient-oriented research. Can J Pain 2018; 2: 191–204.

36 Fitzcharles M-A, Brachaniec M, Cooper L, et al. A paradigm change to inform fibromyalgia research priorities by engaging patients and health care professionals. Can J Pain 2017; 1: 137–47.

37 Harris PA, Taylor R, Thielke R, Payne J, Gonzalez N, Conde JG. Research electronic data capture (REDCap)—A metadata-driven methodology and workflow process for providing translational research informatics support. J Biomed Inform 2009; 42: 377–81.

38 Harris PA, Taylor R, Minor BL, et al. The REDCap Consortium: Building an International Community of Software Platform Partners. J Biomed Inform 2019; 95: 103208.

39 Andersen LN, Kristensen KL, Howell CM, Rathleff MS, Fonager K, Lyng KD. What matters to people with chronic musculoskeletal pain consulting general practice? Comparing research priorities across different sectors. Scand J Pain 2023; 23: 759–66.

40 Braun V, Clarke V. Using thematic analysis in psychology. Qualitative Research in Psychology 2006; : 77–101.

41 Levelink M, Voigt-Barbarowicz M, Brütt AL. Priorities of patients, caregivers and health- care professionals for health research – A systematic review. Health Expect 2020; 23: 992– 1006.

42 Hansen PB, Bahnsen M, Nørgaard MS, Jepsen JF, Rathleff MS, Lyng KD. Effectiveness of Implementation Interventions in Musculoskeletal Healthcare: A Systematic Review. medRxiv 2023; : 2023.11.29.23299209.

43 Straus SE, Tetroe J, Graham I. Defining knowledge translation. Can Méd Assoc J 2009; 181: 165–8.

44 WHO. (2017b). EVIPNet. http://www.euro.who.int/en/data-and-evidence/evidence-informed-policy-making/evidence-informed-policy-network-evipnet.

45 Dubé M-O, Desmeules F, Lewis JS, Roy J-S. Does the addition of motor control or strengthening exercises to education result in better outcomes for rotator cuff-related shoulder pain? A multiarm randomised controlled trial. Br J Sports Med 2023; 57: 457–63.

46 Zadro JR, Ferreira GE, Muller R, et al. Education can reassure people with rotator cuff– related shoulder pain: a 3-arm, randomised, online experiment. PAIN 2024; 165: 951–8.

47 Lapner P, Henry P, Athwal GS, et al. Treatment of rotator cuff tears: a systematic review and meta-analysis. J Shoulder Elb Surg 2022; 31: e120–9.

48 Challoumas D, Biddle M, McLean M, Millar NL. Comparison of Treatments for Frozen Shoulder. Jama Netw Open 2020; 3: e2029581.

49 Singh JA, Sperling J, Buchbinder R, McMaken K. Surgery for shoulder osteoarthritis. Cochrane Db Syst Rev 2010; : CD008089.

50 Glazebrook H, Miller B, Wong I. Anterior Shoulder Instability: A Systematic Review of the Quality and Quantity of the Current Literature for Surgical Treatment. Orthop J Sports Medicine 2018; 6: 2325967118805983.

51 Pieters L, Lewis J, Kuppens K, et al. An Update of Systematic Reviews Examining the Effectiveness of Conservative Physical Therapy Interventions for Subacromial Shoulder Pain. J Orthop Sports Phys Ther 2020; 50: 131–41.

52 Griffin J, Jaggi A, Daniell H, Chester R. A systematic review to compare physiotherapy treatment programmes for atraumatic shoulder instability. Shoulder Elb 2022; : 175857322210807.

53 Vandvik PO, Lähdeoja T, Ardern C, et al. Subacromial decompression surgery for adults with shoulder pain: a clinical practice guideline. Bmj 2019; 364: l294.

54 Lafrance S, Charron M, Roy J-S, et al. Diagnosing, Managing, and Supporting Return to Work of Adults With Rotator Cuff Disorders: A Clinical Practice Guideline. J Orthop Sport Phys 2022; 52: 647–64.

55 Curtis DM, Bradley AT, Lin Y, et al. National Trends Show Declining Use of Arthroscopic Subacromial Decompression Without Rotator Cuff Repair. Arthrosc: J Arthrosc Relat Surg 2021; 37: 3397–404.

56 Ozdag Y, Hayes DS, Garcia VC, et al. Surgeon Factors and Trends Associated With the Use of Subacromial Decompression at the Time of Rotator Cuff Repair. J Hand Surg 2024; 49: 465–71.

57 Lyng KD, Djurtoft C, Bruun MK, et al. What is known and what is still unknown within chronic musculoskeletal pain? A systematic evidence and gap map. Pain 2023; Publish Ahead of Print. DOI:10.1097/j.pain.0000000000002855.

58 Deering K, Brimblecombe N, Matonhodze JC, Nolan F, Collins DA, Renwick L. Methodological procedures for priority setting mental health research: a systematic review summarising the methods, designs and frameworks involved with priority setting. Heal Res Polic Syst 2023; 21: 64.

