## Supplementary figures and images for "Shouldering our Way into a More Meaningful Research Agenda for Atraumatic Shoulder Pain: *A Priority Setting Study*"

### Figure 1

#### FIGURE 1. OVERVIEW OF STUDY PHASES 1-6

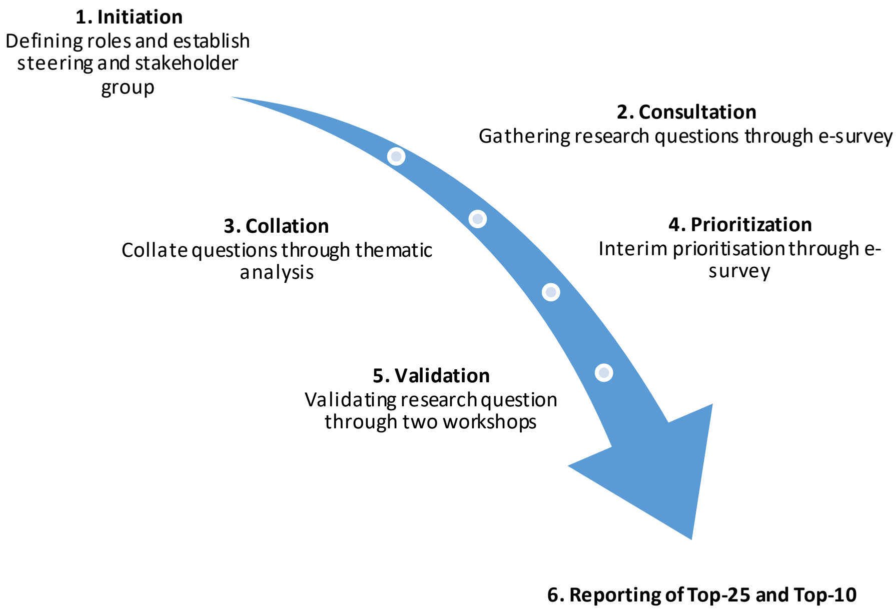
