## Supplementary material for "Shouldering our Way into a More Meaningful Research Agenda for Atraumatic Shoulder Pain: *A Priority Setting Study*": Tables and panel

### Table 1. Participants' Characteristics

| | No. (%) of participants<br>Total n = 608 | | | |
|---|---|---|---|---|
| | Collecting research questions<br>n = 297 (48.8%) | Interim Prioritisation<br>n = 290 (47.6%) | Workshop 1<br>n = 11 (1.8%) | Workshop 2<br>n = 10 (1.6%) |
| **Table 1. Participant Characteristics** | | | | |
| **Participants (% of sub-sample)** | | | | |
| Person living with chronic MSK pain | **230 (77.4)** | **147 (50.6)** | - | **8 (80)** |
| Career to a person living with chronic MSK pain | **1 (0.3)** | **8 (2.7)** | - | **2 (20)** |
| Medical Doctor | **20 (6.7)** | **25 (8.6)** | **4 (36.3)** | - |
| Physiotherapist | **30 (10.1)** | **97 (33.4)** | **3 (27.2)** | - |
| Other HCPs (Psychologist, Nurse, Chiropractor, Social worker, Researcher) | **16 (5.3)** | **13 (4.4)** | **4 (36.3)** | - |
| **Sex** | | | | |
| Female | **177 (59.5)** | **147 (50.7)** | **6 (54.5)** | **4 (40)** |
| Male | **120 (40.5)** | **143 (49.3)** | **5 (45.4)** | **6 (60)** |
| **Age** | | | | |
| 18-30 | **26 (8.7)** | **51 (17.5)** | **1 (9.0)** | **1 (10)** |
| 31-40 | **65 (21.8)** | **57 (19.6)** | **5 (54.5)** | **2 (20)** |
| 41-50 | **83 (27.9)** | **81 (27.9)** | **3 (27.2)** | **3 (30)** |
| 51-60 | **79 (26.5)** | **60 (20.6)** | **1 (9.0)** | **2 (20)** |
| 61-70 | **33 (11.1)** | **29 (10)** | **1 (9.0)** | **1 (10)** |
| 71-80 | **3 (1.0)** | **7 (2.4)** | - | **1 (10)** |
| 80+ | **8 (2.6)** | **5 (1.7)** | - | - |
| **Region** | | | | |
| Region of Northern Jutland | **64 (21.5)** | **61 (21.0)** | **2 (18.1)** | **2 (20)** |
| Region Central Denmark | **60 (20.2)** | **93 (32.0)** | **2 (18.1)** | **1 (10)** |
| Region Southern Denmark | **58 (19.5)** | **51 (17.5)** | **2 (18.1)** | **3 (30)** |

| | | | | |
|---|---|---|---|---|
| Region Zealand | **56 (18.5)** | **22 (7.5)** | **2 (18.1)** | **3 (30)** |
| Region Capital | **59 (19.8)** | **58 (20)** | **3 (27.2)** | **1 (10)** |
| **Ethnicity** | | | | |
| Danish | **250 (84.1)** | **266 (91.7)** | **9 (81.8)** | **3 (30)** |
| Immigrant, non-western | **12 (4.0)** | **5 (1.7)** | **1 (9.0)** | **1 (10)** |
| Immigrant, western | **2 (0.6)** | - | **1 (9.0)** | **2 (20)** |
| Descendant, non-western | **1 (0.3)** | - | - | **2 (20)** |
| Descendant, western | **18 (6.0)** | **2 (0.6)** | - | **2 (20)** |
| Do not want to state | **14 (4.7)** | **17 (5.8)** | - | - |
| **Work Situation (People living with Atraumatic Shoulder Pain)** | | | | |
| Student | **18 (6.0)** | **37 (12.7)** | - | **2 (20)** |
| Full-time employed | **177 (59.5)** | **165 (56.8)** | **10 (91)** | **5 (50)** |
| Part-time employed | **36 (12.1)** | **41 (14.1)** | **1 (9.0)** | **1 (10)** |
| Sick Leave | **51 (17.1)** | **32 (11.0)** | - | **2 (20)** |
| Retired/early retirement/or similar | **12 (4.0)** | **5 (1.7)** | - | - |
| Unemployed | **3 (1.0)** | **10 (3.4)** | - | - |
| **Education (People living with Atraumatic Shoulder Pain)** | | | | |
| No vocotional education | **16 (5.3)** | **10 (3.4)** | - | **2 (20)** |
| One or more short courses | **18 (6.0)** | **2 (0.6)** | - | - |
| Short education (Under 1 year) | **5 (1.6)** | **2 (0.6)** | - | - |
| Vocational education (e.g., Hairdresser, Secretary, Craftsman) | **65 (21.8)** | **68 (23.4)** | - | **2 (20)** |
| Short Academy Profession Degree (e.g., Police, Multimedia-designer) | **30 (10.1)** | **25 (8.6)** | - | **2 (20)** |
| Intermediate Academy Profession Degree (e.g., Nurse, Physiotherapist, Teacher) | **70 (23.5)** | **25 (8.6)** | - | **1 (10)** |
| Long Academy Profession Degree (Medical Doctor, Pscyhologist, Engineer) | **36 (12.1)** | **16 (5.5)** | - | **1 (10)** |

### Table 2. Main themes derived from consultation and collation

| Main themes derived from consultation and collation | |
|---|---|
| **Aids**<br>Knowledge of which aids are available and the effect of these aids.<br><br>**Diagnosis**<br>Knowledge of how we can make a diagnosis more precisely and faster for the individual.<br><br>**Economy**<br>Knowledge of how shoulder pain affects finances for the individual and society.<br><br>**Education**<br>Knowledge of how more knowledge about shoulder pain can improve patients' progress.<br><br>**End-user Involvement**<br>Knowledge of how patients can feel more heard by the health professionals and become more involved in their own process.<br><br>**Individual**<br>Knowledge of how shoulder pain affects the individual's quality of life and time with family.<br><br>**Imaging**<br>Knowledge of the role of imaging (e.g., MR scanner, ultrasound) in the treatment of shoulder pain.<br><br>**Mechanism of Action**<br>Knowledge of which mechanisms cause shoulder pain. Prevention - Knowledge of how we can prevent shoulder pain. | **Patient Journey**<br>Knowledge of how the patient process can be improved and made more efficient and how patients can avoid being stigmatised.<br><br>**Prognosis**<br>Knowledge of how shoulder pain develops and when you can expect improvement/disappearance.<br><br>**Research**<br>Knowledge of how healthcare professionals can be up to date on the latest research and how patients with shoulder pain can help research.<br><br>**Risk Factors**<br>Knowledge of which factors put people at risk of developing shoulder pain<br><br>**Self-management**<br>Knowledge of how patients can best manage their own pain.<br><br>**Sleep**<br>Knowledge of how sleep affects shoulder pain.<br><br>**Treatment**<br>Knowledge of how we best treat shoulder pain.<br><br>**Work**<br>Knowledge of how the job situation affects shoulder pain and how can work be adapted to one's shoulder pain? |

### Table 3. Top 25 Research Questions based on Interim Prioritization

| No. | Research Question | Scoring | Corresponding Main Theme |
|---|---|---|---|
| 1. | Which exercise regimen is the most effective for treating people with atraumatic shoulder pain? | 95 | Treatment |
| 2. | How can people with atraumatic shoulder pain learn to manage their own pain, and which strategies can facilitate this process? | 88 | Self-management |
| 3. | How effective is painful exercising compared to non-painful exercising for treating people with atraumatic shoulder pain? | 71 | Treatment |
| 4. | How can atraumatic shoulder pain be prevented? | 69 | Treatment |
| 5. | Which exercise dose is the most effective for treating people with atraumatic shoulder pain? | 69 | Treatment |
| 6. | Who benefits from surgery, and who doesn't? | 63 | Treatment |
| 7. | How can the return-to-previous function be predicted? | 62 | Prognosis |
| 8. | What is the effectiveness of passive modalities such as (ultrasound, manual therapy etc.)? | 60 | Treatment |
| 9. | What is the risk of developing post-operative pain, and who develops it? | 56 | Risk Factors |
| 10. | Which mechanism is responsible for developing atraumatic shoulder pain? | 53 | Mechanism of Action |
| 11. | How can return-to-previous activities be predicted? | 53 | Prognosis |
| 12. | What is the effectiveness of load management for treating people with atraumatic shoulder pain? | 53 | Treatment |
| 13. | How can we translate evidence into practice? | 51 | Research |
| 14. | How do biological changes (e.g., inflammation, previous injuries, hormones) influence the lived experience? | 49 | Mechanism of Action |
| 15. | Which coping strategies support people with atraumatic shoulder pain in managing their pain? | 48 | Self-management |
| 16. | What is the effectiveness of shared decision-making, and how can it be ensured that the choice of treatments has been decided based on the alliance between people with atraumatic shoulder pain and healthcare practitioners? | 47 | End-user Involvement |
| 17. | What is the effectiveness of self-management compared to supervised/group-based rehabilitation? | 47 | Treatment |
| 18. | How much of improvements in health over time can be attributed to natural healing processes? | 46 | Mechanism of Action |
| 19. | How does atraumatic shoulder pain influence quality of life (including family life and mental health)? | 44 | Mechanism of Action |
| 20. | How can a return to work be predicted? | 43 | Work |
| 21. | How does job adaptability and flexibility influence the prognosis for people with atraumatic shoulder pain? | 41 | Work, Prognosis |
| 22. | How can people with atraumatic shoulder pain and healthcare practitioners be educated to ensure the best management of NTSP? | 37 | Education |
| 23. | What is the mechanism for how impaired sleep can worsen atraumatic shoulder pain symptoms? | 35 | Sleep, Mechanism of Action |
| 24. | How can a good therapeutic alliance be created, and how important is it to the patient journey? | 35 | Patient Journey |
| 25. | What is the effectiveness of patient education for managing atraumatic shoulder pain? | 35 | Education |

*** Score indicates the number of times the questions have been rated very important or important**

### Table 4. Top 10 Prioritized Research Questions

| | Top 10 Research Priorities for Atraumatic Shoulder Pain | Voting 1<br>Place on Top-25 | Voting 2<br>Workshop 1 (HCP) | Voting 3<br>Workshop 2 (ATSP and careers) | Voting 4<br>Vote Total* | Corresponding Theme |
|---|---|---|---|---|---|---|
| 1 | How can we improve the translation of research into clinical practice? | 13 | 5 | 4 | 9 | Research |
| 2 | How can we prevent atraumatic shoulder pain? | 4 | 4 | 4 | 8 | Treatment |
| 3 | Who benefits from surgery, and who does not? | 6 | 4 | 2 | 6 | Treatment |
| 4 | Which exercise type and dose is the most effective?† | 1 and 5 | 4 | 2 | 6 | Treatment |
| 5 | How do patients learn to self-manage their pain, and how can it be facilitated? | 2 | 3 | 4 | 7 | Self-management |
| 6 | Which mechanism(s) is responsible for the development? | 10 | 3 | 3 | 6 | Mechanism of Action |
| 7 | What is the effect of patient education? | 25 | 1 | 3 | 4 | Education |
| 8 | What is the effect of self-management compared to supervised treatment? | 17 | 1 | 2 | 3 | Self-management |
| 9 | What is the effectiveness of exercise with pain compared to without pain? | 3 | 1 | 1 | 2 | Treatment |
| 10 | Which coping strategies support patients in managing their pain? | 15 | 1 | 0 | 1 | Self-management |

**W = workshop, HCP = healthcare practitioners, ATSP = people with atraumatic shoulder pain**

***Votes total is based on the sum of the rating from workshop 1 and 2.**

**† This question were collated based on responses from workshop 1 and 2**
